# Epirubicin for the Treatment of Sepsis and Septic Shock (EPOS-1) - a randomized, placebo-controlled phase IIa dose escalation trial targeting disease tolerance to infection

**DOI:** 10.64898/2026.08.23.26360940

**Authors:** Sebastian Weis, Luís Ferreira Moita, Daniel Thomas-Rüddel, Peter Schlattmann, Christiane Helbig, Thomas Lehmann, Patrick Meybohm, Sven-Olaf Kuhn, Tim Rahmel, Heiko Schenk, Thomas Köcher, Brendan Tibbs, Tiago Velho, Johannes Roth, Frank Brunkhorst, Markus H. Gräler, Ralf Claus, Johannes Ehler, Michael Bauer, the EPOS-1 study group, SepNet Critical Care Trials Group collaborators

## Abstract

**Importance:** Pharmacological targeting of host mechanisms that limit sepsis-induced organ dysfunction represents a new therapeutic approach. Preclinical studies showed that low-dose epirubicin enhances tissue damage control and attenuates sepsis severity independently of pathogen burden, thereby promoting disease tolerance to infection. Yet epirubicin can cause myelotoxicity when used in cancer therapy.

**Objective:** To investigate whether low-dose epirubicin can safely be administered to patients with sepsis and septic shock.

**Design, Setting, and Participants:** A randomized, double-blind, placebo-controlled clinical trial conducted in five German University hospitals. Patients with sepsis, defined by Sepsis-3 criteria, were eligible within 48 hours after diagnosis. The first patient was enrolled on October 19, 2022, and the last follow-up was conducted on May 21, 2025.

**Interventions:** Eligible patients were randomized in a 4:1 ratio to receive either placebo or low-dose epirubicin in addition to standard care. There were three consecutive phases. Patients in the epirubicin group received a single dose of epirubicin (either 3.75 mg/m^2^, 7.5 mg/m^2^ or 15 mg/m^2^, depending on study phase).

**Main Outcomes and Measures:** The primary endpoint of the trial was the 14-day myelotoxicity. Secondary and explorative outcomes included 90-day mortality, the degree of organ dysfunction as assessed by SOFA score, PK/PD modelling and cytokine release.

**Results:** Of 854 patients assessed for eligibility, 32 were randomized and 31 were included in the primary analysis population. Six participants received placebo, nine participants received 3.75 mg/m^2^, nine received 7.5 mg/m^2^ and eight individuals received 15 mg/m^2^ epirubicin, respectively. There was no myelotoxicity in any group. Mortality at 90 days and SOFA-scores were not significantly different between groups. Two of 39 SAEs in the epirubicin group were assessed by the investigators as possibly related to epirubicin,

**Conclusions and Relevance:** Among patients with sepsis and septic shock, low dose epirubicin was not associated with increased myelotoxicity.

## INTRODUCTION

The host’s defense against infection is comprised of two complementary strategies: resistance, which reduces pathogen burden, and disease tolerance, which preserves host fitness by limiting tissue injury, maintaining organ function, and promoting tissue repair without directly affecting pathogen clearance ^1^. Whereas in the clinical routine resistance mechanisms are already augmented through antimicrobial therapy, vaccination, and source control, disease tolerance mechanisms have remained largely unexplored as a therapeutic target ^2^. Experimental studies have shown that disease tolerance is controlled by evolutionarily conserved cellular stress and damage response pathways that protect tissues from infection-associated injury while sustaining organ function. ^3–7^. This aspect becomes particularly relevant in sepsis, in which organ dysfunction is the defining feature of disease severity and the principal determinant of mortality ^8^. Recent observational work could demonstrate that molecular signatures reflecting host resistance, disease tolerance, and tissue damage are associated with clinical outcomes in sepsis patients ^9^.

Anthracyclines have recently emerged as candidate pharmacological inducers of disease tolerance. In experimental models of sepsis, low-dose epirubicin improved survival without reducing pathogen burden, indicating that its protective effects were mediated through enhancement of disease tolerance rather than antimicrobial activity ^10^. The proposed mechanism of action differs fundamentally from conventional antimicrobial therapies. Low-dose anthracyclines induce adaptive DNA damage response pathways, including activation of ataxia telangiectasia mutated (ATM) kinase, thereby enhancing cellular stress resistance and limiting tissue injury ^10^. Similar protective effects have subsequently been described for other modulators of DNA damage responses, including topoisomerase I inhibitors ^11^ suggesting that pharmacologic activation of cellular stress-response pathways may represent a broader therapeutic strategy for acute severe infection. Importantly, these benefits were observed even when treatment was initiated 24 hours after sepsis onset, supporting the translational potential of this approach ^10^. Epirubicin has been used in clinical practice for decades and has an established safety profile at substantially higher doses in oncology ^12^. However, its safety in patients with sepsis has not previously been investigated We therefore conducted a randomized, placebo-controlled, multicenter phase II trial to evaluate the safety and feasibility of low-dose epirubicin in patients with sepsis or septic shock as the first clinical investigation of pharmacological enhancement of disease tolerance during infection.

## METHODS AND MATERIALS

### Trial design, participants and intervention

EPOS-1 was a prospective, randomized, double-blind, placebo-controlled, dose-escalation phase IIa trial conducted at five German centers. Adults (aged ≥18 years) with sepsis admitted to intensive care units (ICUs) or intermediate care units (IMCs) were randomized to receive a single intravenous infusion of epirubicin or placebo in addition to standard of care. Detailed inclusion and exclusion criteria are provided in *Supplement (Suppl.) Table 1*.

**Table 1:** Patient characteristics - demographics, admission type and comorbidities. Age is summarized by mean and standard deviation, while absolute and relative frequencies are reported for all categorical variables.

|  | Placebo<br>No. (%)<br>(n=6) | Phase I<br>3.75 mg/m<sup>2</sup><br>No. (%)<br>(n=9) | Phase II<br>7.5 mg/m<sup>2</sup><br>No. (%)<br>(n=9) | Phase III<br>15 mg/m<sup>2</sup><br>No. (%)<br>(n=8) | Epirubicin all<br>patients<br>No. (%)<br>(n=26) |
| --- | --- | --- | --- | --- | --- |
| Age (years±SD) | 67.3±16.3 | 64.2±11.8 | 67.9±11.2 | 64.3±15.3 | 65.5±12.4 |
| Female gender | 1 (16.7) | 1 (11.1) | 0 (0) | 3 (37.5) | 4 (15.6) |
| Type of admission |  |  |  |  |  |
| Scheduled surgical | 3 (50.0) | 3 (33.3) | 2 (22.2) | 0 (0) | 5 (19.2) |
| Medical | 1 (16.7) | 4 (44.4) | 5 (55.6) | 2 (25.0) | 11 (42.3) |
| Unscheduled surgical | 2 (33.3) | 2 (22.2) | 2 (22.2) | 6 (75.0) | 10 (38.5) |
| APACHE II score (mean, SD) |  |  |  |  |  |
| SAPS 2 score (mean, SD) | 38.5 (10.6) | 49.2 (16.2) | 44.1 (16.1) | 47.8 (10.8) | 47.1 (14.2) |
| qSOFA score (mean, SD) | 2.2 (0.8) | 2 (0.9) | 2.2 (0.4) | 2 (0.9) | 2.1 (0.7) |
| SOFA score (mean, SD) | 7 (1.4) | 8.9 (3.3) | 8.9 (3.0) | 8.6 (1.8) | 8.8 (2.7) |
| Required mechanical ventilation No (%) | 4 (66.7) | 6 (66.7) | 5 (55.6) | 6 (75) | 17 (65.4) |
| *Preexisting conditions* |  |  |  |  |  |
| Myocardial infarction | 1 (16.7) | 0 (0) | 2 (22.2) | 0 (0) | 2 (7.7) |
| Congestive Heart failure | 2 (33.3) | 2 (22.2) | 6 (66.7) | 2 (25) | 10 (38.5) |
| Peripheral Vascular Disease | 2 (33.3) | 1 (11.1) | 2 (22.2) | 1 (12.5) | 4 (15.4) |
| Cerebrovascular Disease | 0 (0) | 0 (0) | 1 (11.1) | 0 (0) | 1 (3.8) |
| Dementia | 0 (0) | 0 (0) | 0 (0) | 0 (0) | 0 (0) |
| Chronic pulmonary disease | 1 (16.7) | 1 (11.1) | 1 (11.1) | 0 (0) | 2 (7.7) |
| Connective tissue disease | 0 (0) | 0 (0) | 0 (0) | 0 (0) | 0 (0) |
| Peptic ulcer | 1 (16.7) | 0 (0) | 0 (0) | 0 (0) | 0 (0) |
| Mild Liver Disease | 0 (0) | 0 (0) | 2 (22.2) | 0 (0) | 2 (7.7) |
| Diabetes | 0 (0) | 3 (33.3) | 4 (44.4) | 4 (50) | 11 (42.3) |
| Paraplegia and Hemiplegia | 0 (0) | 1 (11.1) | 0 (0) | 0 (0) | 1 (3.8) |
| Moderate or severe renal disease | 0 (0) | 1 (11.1) | 4 (44.4) | 0 (0) | 5 (19.2) |
| Diabetes with end organ damage | 1 (16.7) | 1 (11.1) | 3 (33.3) | 0 (0) | 4 (15.4) |
| Any tumor | 0 (0) | 1 (11.1) | 2 (22.2) | 1 (12.5) | 4 (15.4) |
| Leukemia or Lymphoma | 0 (0) | 0 (0) | 0 (0) | 0 (0) | 0 (0) |
| Moderate or severe liver disease | 0 (0) | 0 (0) | 0 (0) | 2 (25) | 2 (7.7) |
| Metastatic solid tumor | 1 (16.7) | 0 (0) | 0 (0) | 0 (0) | 0 (0) |
| AIDS | 0 (0) | 0 (0) | 0 (0) | 0 (0) | 0 (0) |

Patients assigned to the intervention received a single intravenous infusion of epirubicin or placebo over 15 minutes. The trial comprised three sequential dose-escalation phases evaluating epirubicin dosages of 3.75, 7.5, and 15 mg/m². The first dosage of 3.75 mg/m² corresponded to 25% of the efficacious dose in the preclinical mouse model and approximately 4% of the dose administered during a standard chemotherapy cycle. After phase I and phase II, recruitment was paused until an independent Data Safety Monitoring Board (DSMB) reviewed the safety data and recommended either continuation to the next dose level or study termination. Following DSMB approval, the dose was escalated to 7.5 mg/m² and subsequently to 15 mg/m², corresponding to the dose shown to be effective in mice and approximately 16% of a standard chemotherapy dose ^10^.

For each dose level, at least eight patients receiving epirubicin and two patients receiving placebo were required to complete the 14-day follow-up before data cleaning, independent statistical analysis, and DSMB review. Due to the mandatory case numbers of eight epirubicin and two placebo patients with completed 14-day follow-up, patients dropping out until day 14 or patients with premature termination of treatment/infusion or known underdosing were replaced and randomization was continued until the mandatory case numbers were reached.

The German Self-Help Group „Deutsche Sepsis-Hilfe e.V“ endorsed the EPOS-1 study.

### Randomization and Allocation

Patient randomization and assignment to the respective group was retrieved by each local pharmacy using of a validated electronic tool (PaRANDies) provided by the Center for Clinical Studies Jena (ZKS). The randomization list was prepared by an independent statistician via a computer-based algorithm and is stratified by study center. For each study phase patients are randomized to one of the two study arms (epirubicin or placebo) in a 4:1 ratio. Study medication/infusion was prepared in the hospital pharmacies of the trial sites by unblinded personnel and then delivered to the ICU/IMC in colored infusion systems to avoid unblinding by the typical red color of epirubicin. Treating physicians, the study team members and the participant were blinded.

### Endpoints

The primary endpoint of the study was safety as assessed by myelotoxicity until day 14. Assessing myelotoxicity in sepsis patients can be complicated since leukopenia, neutropenia and thrombocytopenia are common in sepsis ^13–16^. To differentiate the best possible way between sepsis-associated alterations and epirubicin-induced myelotoxicity the primary safety endpoint was defined as follows: neutropenia of grade 3 or 4 at two consecutive study visits up to day 14 or thrombocytopenia of grade 3 or 4 at two consecutive study days up to day 14 accompanied by neutropenia or thrombocytopenia of grade 2, 3 or 4 at both study days and accompanied by an immature platelet fraction (IPF) below 2.5% at one or two of the consecutive study visits (*Suppl. Figure 1, Suppl. Table 2*). Secondary endpoints for safety were cardiotoxicity, assessed by left ventricular ejection fraction (LVEF) carried out by transthoracic echocardiography (TTE) 7 days after epirubicin administration, the frequency of other typical side effects, and the overall rate of adverse and serious adverse events. The inflammatory response was determined by measuring procalcitonin-(PCT), and C-reactive protein-(CRP) serum levels. A “success” rate was defined as a decrease of PCT serum concentrations by 80% or more of its intra-individual peak value or to 0.5 μg/L or lower within 72 hours after randomization (following the “Stop Antibiotics on Procalcitonin guidance Study” (SAPS) by de Jong *et al.* ^17^ . Daily changes in SOFA scores and need for organ support therapies over time were documented. Mortality was assessed at days 14, 28, and 90 after randomization. Quality of life was assessed at day 90 in survivors by Short Form 36 Health Questionnaire (SF-36). Explorative analyses included pharmacokinetics and pharmacodynamics of epirubicin, cytokine release and determination of DNA damage in leukocytes by measurement of phosphorylated H2A histones (pH2AX).

### Sample Size Calculation

EPOS-1 was an exploratory trial to test the safety of low-dose epirubicin administration in patients with sepsis for the first time. Sample size calculations were based on data from cancer patients, who receive epirubicin doses four times higher. Myelotoxicity was observed in cancer patients that received repetitive courses of epirubicin. Herait *et al.* reported grade 3 to 4 leucopenia in approximately 20% of patients that were treated every 3-4 weeks using a dose of 85 to 90 mg/m^2^ epirubicin ^18^. In another trial, myelotoxicity grade 3 to 4 using the WHO classification was reported in 14 of patients receiving 71-75 mg/m^2^ every 3 weeks ^19^. Based on the assumption that the probability of a myelotoxicity is 18% the probability of observing at least one myelotoxicity out of eight verum-treated patients equals 79.6% based on a binomial distribution. Therefore, a total of 30 patients (8 x verum *vs.* 2 x placebo from each phase) that reached the 14-day safety endpoint was required. Drop-outs until day 14 were replaced until necessary numbers were reached.

### Statistical analysis

The primary analyses at each dose level estimated the proportion of patients experiencing myelotoxicity, together with the corresponding 95% confidence interval based on the method by Clopper-Pearson. The incidences of thrombocytopenia and neutropenia were compared using two-sided Fisher’s exact tests. All other analyses were descriptive. Continuous variables were summarized using the median and 25^th^ and 75^th^ percentiles or mean and standard deviation, whereas categorical variables were presented as absolute and relative frequencies.

Following completion of the first and second dose-escalation phases, the Data Safety Monitoring Board (DSMB) convened to recommend whether the study should be discontinued or whether the next higher dose level should be initiated. The DSMB received the relevant prespecified analyses and raw data. The primary stopping rule was based on increased toxicity in epirubicin groups as assessed by myelotoxicity. For the analysis of mortality initially a logistic regression model with random intercepts for patients was performed, followed by an exact logistic regression using SAS PROC LOGISTIC. Treatment groups Placebo, 3.75 mg/m², 7.5 mg/m² and 15 mg/m² with placebo as reference were investigated. Additionally, time points 14, 28 and 90 days with 14 days as a reference were considered. Epirubicin peak levels, epirubicin area under the curve and levels of phosphorylated H2A histones (pH2AX) where compared usingthe Kruskal-Wallis test with two-stage linear step-up procedure of Benjamini, Krieger and Yekutieli as post-hoc test. Statistical analyses were performed using SAS version 9.4 (SAS Institute Inc., Cary, NC, USA). Exploratory data analyses were conducted using GraphPad Prism version 10.3.1 (GraphPad Software, Boston, MA, USA).

## Results

### Trial participants

Of 854 patients assessed for eligibility, 32 were randomized and 31 were included in the primary analysis population. Patients had a mean age of 65.8 years (standard deviation: 12.9 years), and 5 of 32 (15.6%) patients were female. Six patients were assigned to the placebo group and 26 to the epirubicin group. Nine patients each were recruited in Phase I (3.75 mg/m^2^) and Phase II (7.5 mg/m^2^) of the study in the epirubicin group, and eight patients in Phase III (15 mg/m^2^) (*Figure (Fig.) 1*)). The two groups were comparable with respect to patient characteristics, admission type, sepsis severity and preexisting comorbidities (*Table 1*).

**Figure 1:**
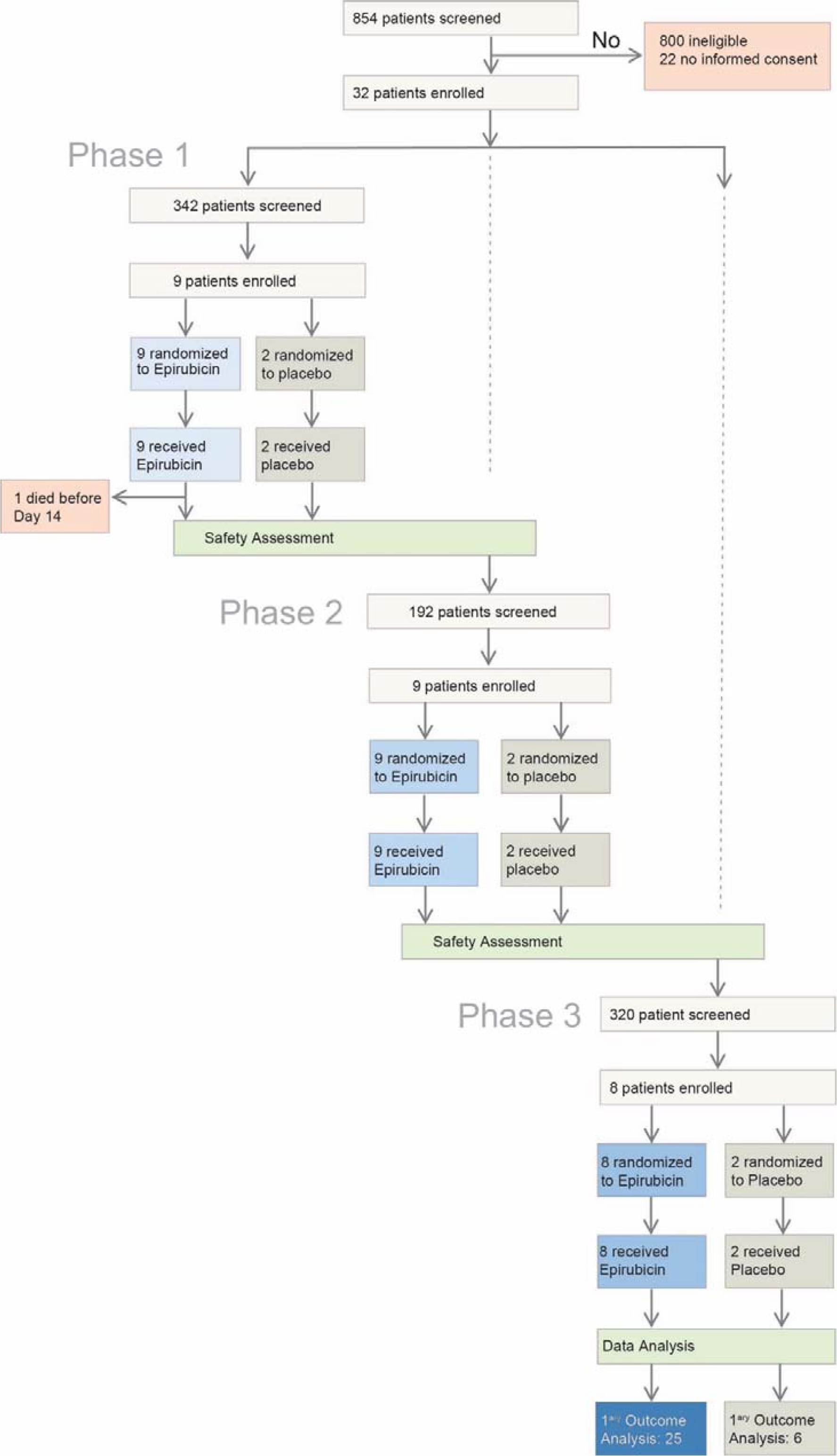
Study flow chart of the EPOS-1 trial.

### Primary End Point

None of the participants developed myelotoxicity until day 14 after the intervention (*Table 2*) (i.e., proportion of myelotoxicity is 0% in both groups in any phase of the trial). There was no case of Grade 2-4 thrombocytopenia in the placebo group. In the epirubicin group seven patients (26.9%) had Grade 1, 2 patients (7.7%) had Grade 2 and one patient (3.8%) had Grade 3 thrombocytopenia. The incidence of thrombocytopenia was not statistically significant different between both groups (p=0.514). No neutropenia occurred in the placebo group, whereas in the epirubicin group one case each (3.1%) of Grade 2 and Grade 3 neutropenia was observed. There was no significant difference between the groups (p=1.000). In the placebo group, three patients (50%) had an immature platelet fraction (IPF) values below 2.5%, compared to 11 patients (42.3%) in the epirubicin group. The difference in IPF rates between groups was not statistically significant (p=1.000). Incidences in the three different phases of the trial is shown in *Table 2*.

**Table 2:** Thrombocytopenia, neutropenia and IPF in intervention (phase I-III) and placebo group, absolute and relative frequencies Neutropenia in intervention (phase I-III) and placebo group, absolute and relative frequencies. Abbreviations: CI - confidence interval; LLN - lower limit of normal; IPF - immature platelet fraction

|  | Placebo | Phase I: 3.75 mg/m<sup>2</sup> | Phase II: 7.5 mg/m<sup>2</sup> | Phase III: 15 mg/m<sup>2</sup> | Epirubicin all patients |
| --- | --- | --- | --- | --- | --- |
| Primary Endpoint | N (95%-CI) | N (95%-CI) | N (95%-CI) | N (95%-CI) | N (95%-CI) |
| Myelotoxicity | 0 | 0 | 0 | 0 | 0 |
| Maximum grade of thrombocytopenia, cumulative | N (%) | N (%) | N (%) | N (%) | N (%) |
| Grade 1 (150,000-75,000/μL) | 6 (100) | 6 (66.7) | 6 (66.7) | 4 (50.0) | 16 (61.5) |
| Grade 2 (<75,000/μL) | 0 | 2 (22.2) | 3 (33.3) | 2 (25.0) | 7 (26.9) |
| Grade 3 (<75,000-50,000/μL) | 0 | 1 (11.1) | 0 | 1 (12.5) | 2 (7.7) |
| Grade 4 (<50,000-25,000/μL) | 0 | 0 | 0 | 1 (12.5) | 1 (3.8) |
| Maximum grade of neutropenia, cumulative | N (%) | N (%) | N (%) | N (%) | N (%) |
| no Neutropenia | 6 (100) | 9 (100.0) | 8 (88.9) | 7 (87.5) | 24 (92.3) |
| Grade 2 (<1,500-1,000/μL) | 0 | 0 | 1 (11.1) | 0 | 1 (3.8) |
| Grade 3 (<1,000-500/μL) | 0 | 0 | 0 | 1 (12.5) | 1 (3.8) |
| IPF < 2.5%, cumulative | N (%) | N (%) | N (%) | N(%) | N (%) |
| IPF ≥ 2.5% | 3 (50) | 6 (66.7) | 4 (44.4) | 5 (62.5) | 15 (57.7) |
| IPF < 2.5% | 3 (50) | 3 (33.3) | 5 (55.6) | 3 (37.5) | 11 (42.3) |

**Table 3:**
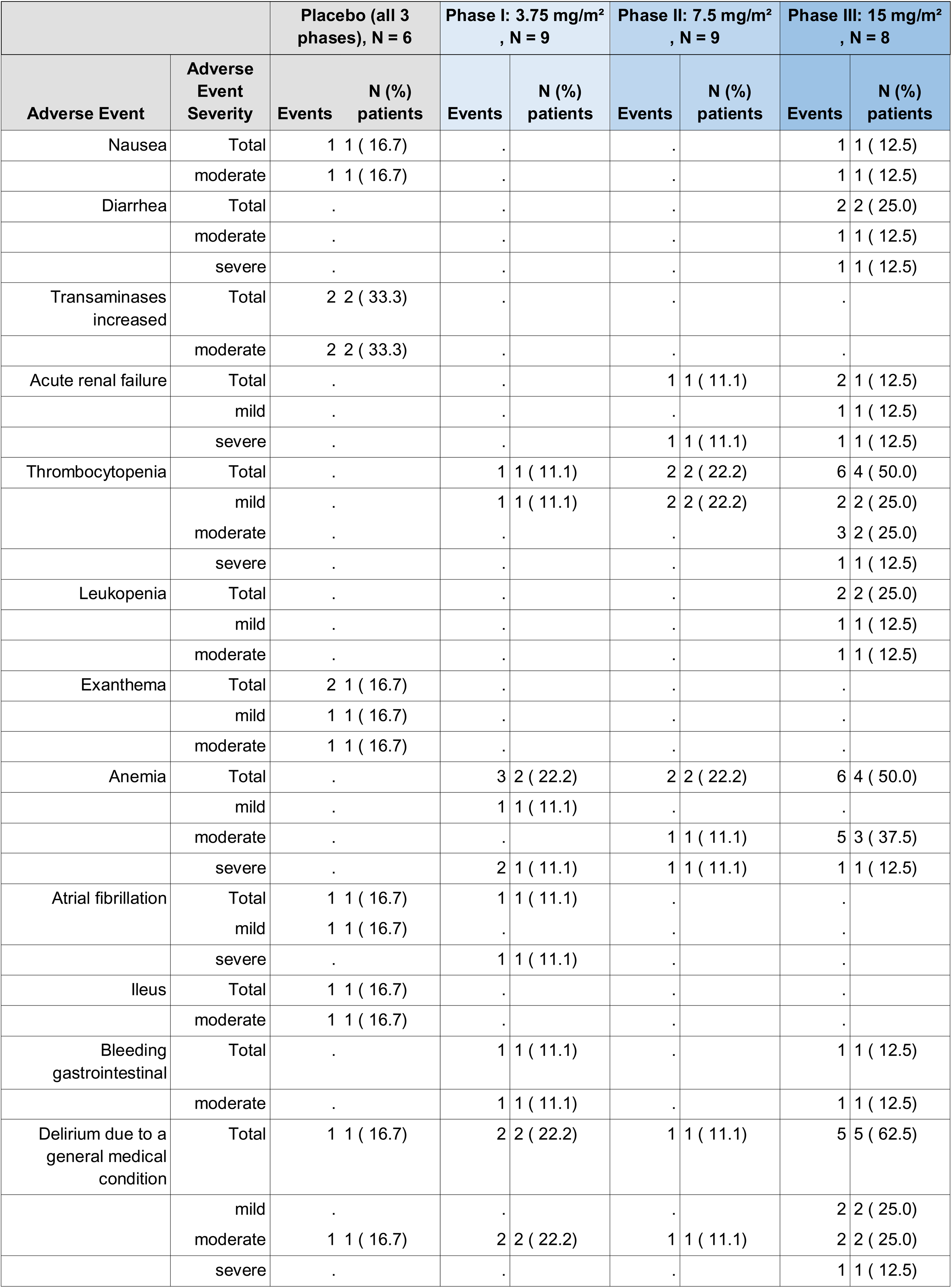
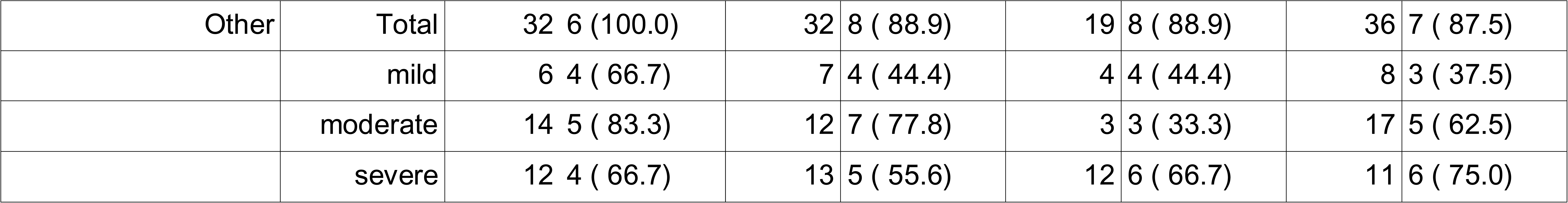
Incidence and severity of AEs (mild/moderate/severe).

### Secondary Outcomes

There were no signs of early cardiotoxicity as assessed by left ventricular ejection fraction at day 7 after study drug application (*Suppl.Fig. 3A*). Until day 90 one of six patients (16.7%) in the placebo group and 5 of 26 patients (19.2%) in the epirubicin group died. Using an exact logistic regression, we found neither an effect of dose (Score = 1.98, p-value= 0.62) or time (Score= 3.28, p-value 0.26). The same was observed when assessing odds ratios based on this model with respect to dose or time (*Suppl. Table 3*). Mortality data on days 14, 28 and 90 in the different epirubicin groups are shown in (*Suppl.Table 4*).

Causes of death of overall six patients in the trial are provided in *Suppl.Table 5*. There were no significant differences in daily SOFA scores, and paO_2_/FiO_2_ ratios from baseline to day 7 between the groups. The number of vasopressor-free days were not different (*Suppl. Fig. 3B*). Daily urine output and fluid balances at days 1-3 were also not different (*Suppl.Fig. 3C-F)*. In addition, epirubicin did not alter markers of systemic inflammation as assessed by PCT-, CRP- and interleukin-6 serum levels (*Suppl.Fig. 4A-D).* Patient-reported quality of life as measured by the Short Form-36 Health Survey (SF-36) at the end of the follow-up was not different between groups (*Suppl.Fig. 4E)*.

### Adverse Events

All but one participants (31/32) experienced at least one adverse event (AE) during the study. In the placebo group, all six patients (100%) experienced a total of 40 AEs. In the epirubicin group, all nine patients (100%) experienced at least one AE during Phase I (40 AEs in total). In Phase II, 25 AEs were reported in eight of nine patients (88.9%), whereas in Phase III, 61 AEs were reported in all eight treated patients (100%). Overall, 14 AEs in the epirubicin group were considered related to the investigational medicinal product (IMP), compared with one treatment-related AE in the placebo group (*Table 2*). Serious adverse events (SAEs) occurred in both treatment groups. In the placebo group, 10 of 40 AEs (25.0%) were classified as serious. In the epirubicin group, 15 of 40 AEs (37.5%) in Phase I, 12 of 25 AEs (48.0%) in Phase II, and 12 of 61 AEs (19.7%) in Phase III were classified as serious, corresponding to 39 SAEs among 126 reported AEs (31.0%) across all treatment phases (*Suppl.Table 6,7*). Two SAEs in the epirubicin group were assessed by the investigators as possibly related to the drug. These consisted of one soft tissue infection and one device-related infection. Both SAEs were not fully resolved (=ongoing) at the 90 day follow up. After study end the outcome was reported as recovered/resolved. There were no cases of alopezia or mucositis reported. All related SAEs were reported as SUSARs. A detailed incidence analysis by severity is provided in the Supplementary Appendix (*Suppl.Table 6,7*).

### Exploratory Analyses

PK/PD analysis demonstrated a dose-dependent pharmacokinetic response across the three dose levels evaluated. Increasing dose was associated with progressively greater pharmacokinetic c effects, with median peak responses 269 (IQR 60-421) pmol/mL for 3.75mg/m^2^ epirubicin, 581.5 pmol/mL (IQR 376-728) for 7.5mg/m^2^ epirubicin and 1.557 pmol/mL; (IQR 1098-1999) for 15 mg/m^2^ epirubicin, respectively. The pharmacokinetic response was maximum after 15 minutes, which also marked the end of the infusion, followed by a steep decline. The exposure-response relationship was consistent with dose-dependent target engagement over the dose range investigated. A significant dose-dependency was also observed when comparing the median of the peak pharmacokinetic responses or the area under the curve (AUC_3.75 mg/m2_=20.796 pmol·min/mL; SE±5.090) vs. (AUC_7.5 mg/m2_=36.987 pmol·min/mL; SE±5.879) vs. (AUC_15 mg/m2_= 63.273 pmol·min/mL; SE±13.383)(p <0.001) (*Figure 2A,B*). Since epirubicin induces DNA damage, we also assessed the levels of DNA damage in circulating immune cells and found a significant increase in pH2AX 6 hours after infusion of 15 mg/m^2^ epirubicin when compared to placebo.

**Figure 2:**
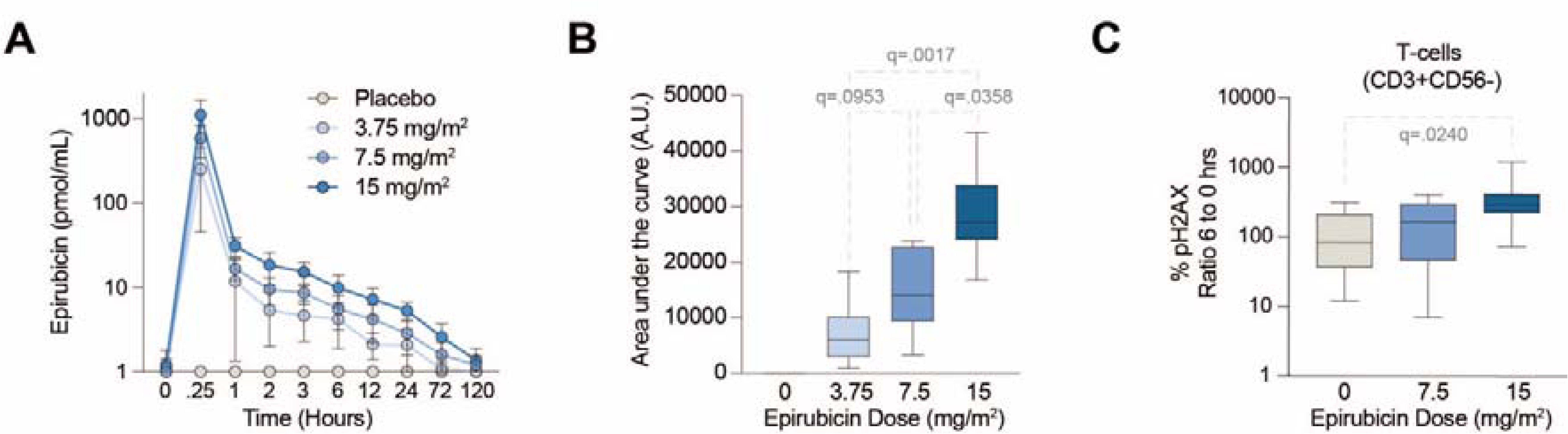
**A**) Plasma epirubicin concentrations over time. Data are presented as mean ± standard deviation. One outlier was removed. All data <1 were set to 1 exclusively for the presentation of the data. **B**) Area under the curve (AUC) of epirubicin plasma levels. **C**) ratio 6 to 0 hours of %pH2AX in CD3+CD56-T-cells. Kruskal-Wallis test with two-stage linear step-up procedure of Benjamini, Krieger and Yekutieli as post-hoc test, in C against placebo. Grey numbers indicate q-value.

## DISCUSSION

In this multicenter phase 2a dose-escalation randomized clinical trial, a single administration of low-dose epirubicin in patients with sepsis was not associated with dose-limiting myelotoxicity or other clinically relevant safety concerns across the three dosage levels evaluated. Beyond establishing the short-term safety of low-dose epirubicin, this study provides the first clinical translation of a host-directed therapeutic strategy promoting disease tolerance to infection ^2^.

The highest dose evaluated in EPOS-1 corresponded to the dose that demonstrated protection in preclinical models ^10^. Epirubicin was selected because it has a more favorable safety profile than doxorubicin while maintaining similar pharmacologic activity differs ^12^. Myelosuppression was chosen as the primary safety endpoint because it represents the principal acute dose-limiting toxicity of anthracyclines ^12,20^. Although previous oncology studies suggested that comparable single doses are generally well tolerated ^21^ this was so far not investigated in critically ill patients with sepsis. The absence of clinically relevant myelotoxicity across all dose levels therefore represents an important translational finding.

Although EPOS-1 was not designed to evaluate efficacy, the absence of major safety signals supports further clinical development of this host-directed therapeutic approach. Future randomized trials should evaluate earlier administration, identify the optimal biological dose, and determine whether pharmacologic induction of disease tolerance reduces organ dysfunction and improves patient-centered outcomes. In addition, incorporation of pharmacodynamic biomarkers may help confirm target engagement and guide dose selection. If efficacy is demonstrated, disease tolerance induction could represent a novel adjunct to standard sepsis therapy

### Limitations

This trial has several limitations. First, the sample size was small, and participants receiving epirubicin were distributed across three dosage levels. Consequently, the study was not powered to detect uncommon adverse events or modest differences in safety between dose groups. Second, because this was an early-phase safety trial, patients at increased risk of myelotoxicity - including those with baseline cytopenias, hematologic disorders, concurrent myelosuppressive therapy, or substantial hepatic or renal dysfunction—were excluded. Therefore, the findings may not be generalizable to all patients with sepsis, particularly those with greater disease severity or comorbidities. Third, study treatment was administered relatively late after sepsis onset. While appropriate for an initial safety assessment, this timing may have reduced the opportunity to observe biological or clinical effects. Finally, the trial was not designed or powered to assess efficacy, precluding conclusions regarding clinical benefit, optimal dose selection, or effect-size estimates for future confirmatory studies.

### Conclusions

In this phase IIa dose-escalation randomized clinical trial, single-dose low-dose epirubicin was not associated with dose-limiting myelotoxicity or other major safety concerns in patients with sepsis. These findings support the feasibility to use assess pharmacologic induction of disease tolerance with epirubicin in humans and provide the rationale for larger randomized trials designed to determine whether induction of disease tolerance as an adjunctive treatment strategy improves clinical outcomes in sepsis.

## Data Availability

All data produced in the present study are available upon reasonable request to the authors

## Contributors

All authors fulfill the ICMJE recommendations for authorships. The roles and contributions of the individual authors are as follows: Sponsor Representative/Principal and coordinating investigator: **SW**; Deputy Coordinating Investigator: **DTR, JE**; Project Manager: **CH**; Protocol writing and planning of the study: **SW, DTR, FB, MB, PS, CH, AH, LFM**; Acquisition of Funding: **SW, FB, TK**. Formulation of initial hypothesis: **SW, LFM**; Trial statistician, sample size calculation and statistical analysis: **TL, PS**; Molecular analysis: **LFM, MHG, RAC, TK**; **BT, TV;** Study Center Coordinators and recruitment of patients**: JE, JR, TR, PM, SOK, HS**. All authors contributed to, read and approved the final manuscript and contributed to the writing.

