## SUPPLEMENT for "Epirubicin for the Treatment of Sepsis and Septic Shock (EPOS-1) - a randomized, placebo-controlled phase IIa dose escalation trial targeting disease tolerance to infection"

#### SUPPLEMENT 1: PROTOCOL MODIFICATIONS

| Substantial Modifications to the study protocol | |
|---|---|
| **Version study protocol** | **Changes/Comments** |
| Version 01 | Version for the initial application to the competent authority. |
| Version 02 | Adaption according to claims from the competent authority (to study protocol version 01). Version for initial application to the ethic committee. |
| **Version 03**<br>10.01.2022 | **Clinical Trial protocol Amendment No.1**<br>Amendment No. 01 belongs to the study protocol version 03 from 10.01.2022. This protocol version replaced study protocol version 02 from 22.10.2021<br><u>Reason for changes</u><br>The study protocol was updated due to additional parameter “leukocyte metabolites” as explorative endpoints. This parameter is measured in PMBC samples in the collaborating laboratory in Vienna. No additional blood sampling is necessary.<br><u>Possible consequences and risks</u><br>The changes in the protocol had no risks for patient safety.<br>Addition of hints served to increase the safety of patients in conduction the trial. |
| **Version 04**<br>07.04.2022 | **Clinical Trial protocol Amendment No.2**<br>Amendment No. 02 belongs to the study protocol version 04 from 07.04.2022. This protocol version replaced study protocol version 03 from 10.01.2022<br><u>Reason for changes</u><br>The study protocol was updated with an extension of the intermediate care (IMC) due to planned admission of one additional trial center (Hannover) that only had an IMC but no intensive care unit (ICU).<br><u>Possible consequences and risks</u><br>The changes in the protocol had no risks for patient safety. |
| **Version 05**<br>13.10.2022 | **Clinical Trial protocol Amendment No.3**<br>Amendment No. 03 belongs to the study protocol version 05 from 13.10.2022. This protocol version replaced study protocol version 04 from 07.04.2022<br><u>Reason for changes</u><br>The study protocol was updated with Section 8.11 - Premature Unblinding to describe processes of premature unblinding of several involved persons in the double-blinded study. In this protocol amendment, the blinded reporting of potential SUSARs to the investigators was concretised.<br><u>Possible consequences and risks</u><br>The changes in the protocol have no risks for patient safety. |
| **Version 06**<br>12.06.2023 | **Clinical Trial protocol Amendment No.4**<br>Amendment No. 04 belongs to the study protocol version 06 from 12.06.2023. This protocol version replaced study protocol version 05 from 13.10.2022.<br><u>Reason for changes</u><br>Planned change in the inclusion and exclusion criteria from which we thought that inclusion rates will raise without increasing the risk of toxicity in the target population. It seemed that the previous inclusion/exclusion criteria (protocol |

| Substantial Modifications to the study protocol | |
|---|---|
| **Version study protocol** | **Changes/Comments** |
| | V05) were too strict to allow the study to be carried out successfully with five centres. Since the primary objective of the EPOS-1 study is to evaluate the safety of epirubicin in sepsis, we had identified two criteria that we believed can be changed:<br>• Extension of the sepsis diagnosis window to 48 hours in order to increase recruitment. While the most rapid application of the drug after diagnosis appears to be useful for an efficacy outcome, it might not be as relevant when assessing safety.<br>• In the case of the mere diagnosis of "active neoplasia" without concomitant chemotherapy, we do not assume an increased risk of toxicity for the patient. Therefore, we deleted this exclusion criterion.<br>Possible consequences and risks<br>The changes in the protocol have no risks for patient safety. |
| **Version 07**<br>04.06.2024 | **Clinical Trial protocol Amendment No.5**<br>Amendment No. 05 belongs to the study protocol version 07 from 04.06.2024. This protocol version replaced study protocol version 06 from 12.06.2023.<br>Reason for changes<br>After a detailed assessment of the 2nd Interim Analysis Report during the regular DSMB meeting the DSMB made the following requests:<br>• The inclusion of an additional blood count measurement after day 14 only for patients that have an unresolved neutropenia/thrombocytopenia at day 14 until this is resolved<br>• The DSMB shall be informed about all neutropenia/thrombocytopenia AEs in a timely manner and will make a corresponding recommendation.<br>Before starting with the Phase 3 (dose escalation to 15 mg/m2 Epirubicin) of the study we amended the protocol with an additional Visit/blood count measurement: latest at Day 28. This visit was used to follow up patients with unresolved Neutropenia or Thrombocytopenia at day 14 (Visit XV) regardless of grade until this event is resolved. This visit could be repeated if necessary. In Phase 3 of the study, the trial sites were instructed to document each Neutropenia grade 1,2,3 or 4 and each or Thrombocytopenia grade 1,2,3 or 4 as an AE and report it to the sponsor via the SAE form. This allowed the sponsor to rapidly forward every cases of neutropenia, thrombocytopenia regardless of grade the DSMB.<br>Possible consequences and risks.<br>The changes in the protocol had no risks for patient safety. The change had no consequence for subjects already treated in the trial. The changes came in to effect with inclusion of patients in the 3rd study phase (dose escalation to 15 mg/m2 Epirubicin). |

**Suppl. Table 1:** Substantial Modifications to the study protocol.

### SUPPLEMENT 2: METHODS AND DATA

#### 2.1 Supplementary Methods

##### Protocol Amendments

There were five amendments to the initial approved study protocol (*Supplement 1*). Due to low inclusion rate in the centers, the causes for screening failures were evaluated and discussed the DSMB. Therefore, the inclusion time window was changed from 24 to 48 hours and the exclusion criteria “active neoplasia” was omitted. This was considered not be associated with an increased risk of toxicity in the target population. In addition, at the second Interim Analysis, the DSMB requested additional blood count measurement after day 14 only for patients that have an unresolved neutropenia/thrombocytopenia at day 14 until this is resolved and that the DSMB shall be informed about all neutropenia/thrombocytopenia AEs in a timely manner. Consequently, in Phase 3, the trial sites were instructed to document each Neutropenia or Thrombocytopenia grade as an AE and report it to the sponsor via the SAE form.

##### Data collection/data management

Data were collected on an electronic case report form (CRF) using OpenClinica® (OpenClinica, LLC, Waltham, MA, USA) by a trained investigator or study assistant at each respective trial center. Monitoring was performed by the ZKS Jena to its local standard operating procedures (SOPs). Monitoring, in general, were performed on-site. All serious adverse events (SAEs), whether related or not related to study medication, had to be reported until 90 days after administration of epirubicin or placebo. Patients or representatives were contacted on day 28 and day 90 after randomization to obtain the survival status of the participants.

##### Further study procedures

Acute physiology data were documented directly before and at seven time points up to 24 hours after the Interventional Medical Product (IMP) administration. Plasma was centrifugated and stored at -80°C for further analysis. Peripheral Blood Monocytic cells (PBMCs) were solated at the trial sited using a commercially available kit (MACSprep™ PBMC Isolation Kit, Miltenyi Biotec) following the manufactures instructions. Plasma cytokines were determined using the LEGENDplex™ human Inflammation Panel 13-plex (BioLegend, San Diego), according to manufacturer instructions.

##### DNA DAMAGE assessment

Peripheral blood was collected from each patient at different timepoints, and PBMCs were isolated using the human MACSprep™ PBMC Isolation Kit (Miltenyi Biotec; Bergish Gladbach, Germany), according to the manufacturer’s instructions. Cell pellets were resuspended in Recovery™ Cell Culture freezing medium (Thermo Fisher Scientific; Massachusetts, USA), placed in a CoolCell® freezing container (Biozym; Hessisch Oldendorf, Germany) and stored at −80°C until analysis. Immediately before analysis cells were thawed in a 37°C water bath, diluted in RPMI 1640 + 10% FBS then washed with PBS + 2% FBS. Cells were stained with anti-CD16 cFluor R668 as a blocking step, followed by remaining cell surface antibodies from Cytek’s eight colour cFluor immunoprofiling kit (R7-40001 Cytek Biosciences, California, USA) to identify lymphocytes, helper T cells, cytotoxic T cells, natural killer (NK) T cells, B cells, NK cells, and non-classical, intermediate, and classical monocytes. Dead cells were excluded by ViaDye Red (Cytek Biosciences, California, USA). Using the Muse® H2A.X Activation Dual Detection Kit (California, USA), cells were then fixed, permeabilised and stained intracellularly for Histone H2A.X and phospho-Histone H2A.X (Ser139). All antibodies and reagents were purchased from Cytek Biosciences (California, USA), recognizing the following antigens: CD45 (HI30), CD3 (SK7), CD4 (SK3), CD8 (SK1), CD19 (HIB19), CD56 (5.1H11), CD16 (3G8), CD14 (M5E2), Histone H2A.X and phospho-Histone H2A.X (Ser139), conjugated to cFluor V547, cFluor V420, cFluor B515, cFluor R780, cFluor V450, cFluor R668, cFluor BYG710, cFluor R720, Alexa Fluor 555 and PE-Cy5. Samples were acquired using a five laser Cytek Aurora spectral cytometer and analysis was performed using FlowJo v10.

#### Epirubicin pharmacokinetics

For epirubicin quantitation a standard curve in citrated plasma (200 µL) obtained from a healthy volunteer ranging from 0.01 to 300 pmol/injection (Epirubicin hydrochloride/European Pharmacopoeia Reference Standard, Taufkirchen, Germany) was prepared, daunorubicin (30 pmoles/injection) was used for normalization, same amount was added to patient samples (200 µL). Following incubation of one hour at room temperature, proteins were precipitated in glass vials by addition of 160 µL isopropanol and 640 µL chloroform (all solvents in Chromasolv LC-MS grade, Honeywell Riedel-de Haën, Seelze, Germany). After vigorous mixing, supernatants were removed following centrifugation (4.500xg, 4 °C, 6 min), evaporated and resolubilized in 100 µL methanol/chloroform mixture (4:1, V/V). Epirubicin analyses were performed in all participants in a randomized manner using liquid chromatography coupled to fluorescence spectrometry. The Shimadzu Prominence HPLC system consisted of an autosampler SIL20A, a DGU20A5R degassing unit, a binary pump LC20AD, and a CTO20AC column oven with a 60 × 2 mm MultoHigh 100 RP 18 column with 3 µm particle size (CS-Chromatographie Service, Langerwehe, Germany), which was maintained at 35 °C. Mobile phase A consisted of 1% (V/V) formic acid in double distilled water, and mobile phase B consisted of 100% methanol. The column was equilibrated in 10% B with a flow rate of 0.4 mL/min. The mobile phase switched to 40% B after sample injection. At 25 min, the mobile phase changed to 10% B, and the flow rate remained constant until the end of the program at 30 minutes. The injection volume per sample was 10 µL; samples were cooled at 8 °C. The HPLC system was coupled to Hitachi L-2480 FL Detector (Xenon lamp; Ex497/Em557 nm, standard bandwidth, PMT Voltage super high, sampling period 200 ms). Retention time of epirubicin was 9.3 min, of daunorubicin 15.0 min with an adequate separation of fluorescent signals. The analytical results (area under curve) were quantified with EZ Chrome Elite for Hitachi (version 3.1.5) based on internal standard samples and an external standard curve as described above.

#### Ethics

This trial was performed in compliance with the International Council for Harmonisation (ICH) guideline on Good Clinical Practice (GCP), the Declaration of Helsinki, and applicable regulatory requirements, including the archiving of essential documents. Personal data included were collected and processed in accordance with the European Union General Data Protection Regulation (GDPR) 679/2016 (EC). The trial was approved by the ethics committee of the Jena University Hospital on 20 December 2021 (2021-2440-AMG-ff) and the German Health Authorities (BfArM) on 08 November 2021. In addition, the local ethics committees at each site approved the study protocol and the study competence of each site. Written informed consent was obtained from all patients or their authorized or legal representatives. If this was not possible before enrolment in due time and the local ethics committee had approved, a deferred consent process where the inability to provide consent and the urgency of participation in the study with possible benefits for the subject must confirmed in writing by an independent physician, and the patient is enrolled without informed consent. As soon as the legal representative of the patient was available, written informed consent was immediately obtained.

#### 2.2 Supplementary Figures

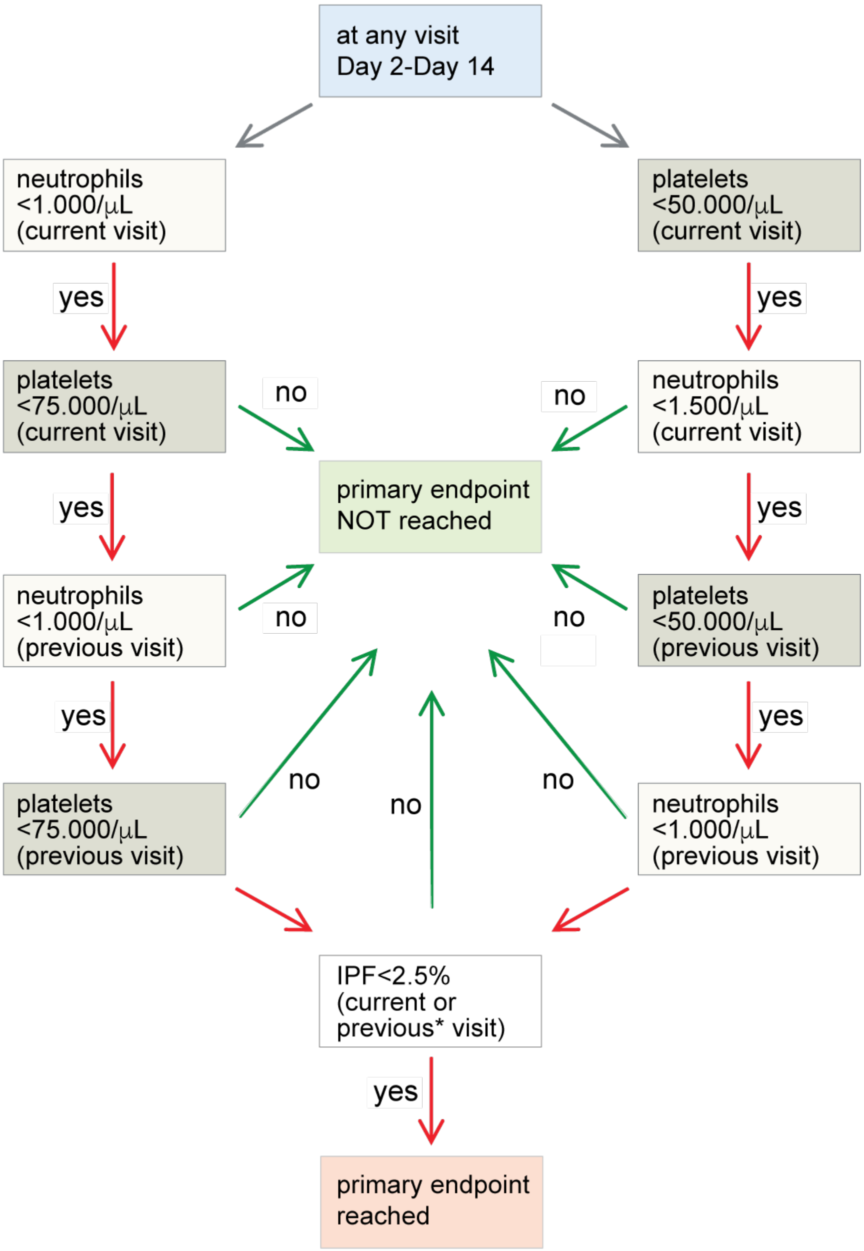


**Suppl.Figure 1:** Flow diagram for the primary endpoint myelotoxicity.

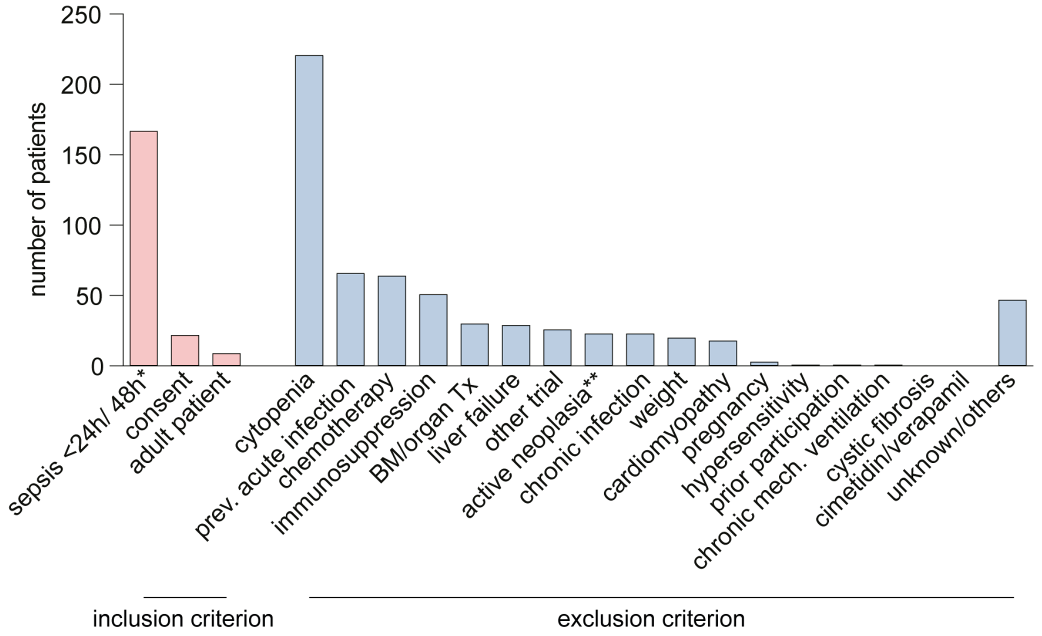


**Suppl.Figure 2**: Reason for exclusion at screening. * changed to 48 hours with 4th protocol amendment. ** omitted with 4th protocol amendment. Cytopenia refers to either thrombocytopenia, leukopenia or thrombopenia.

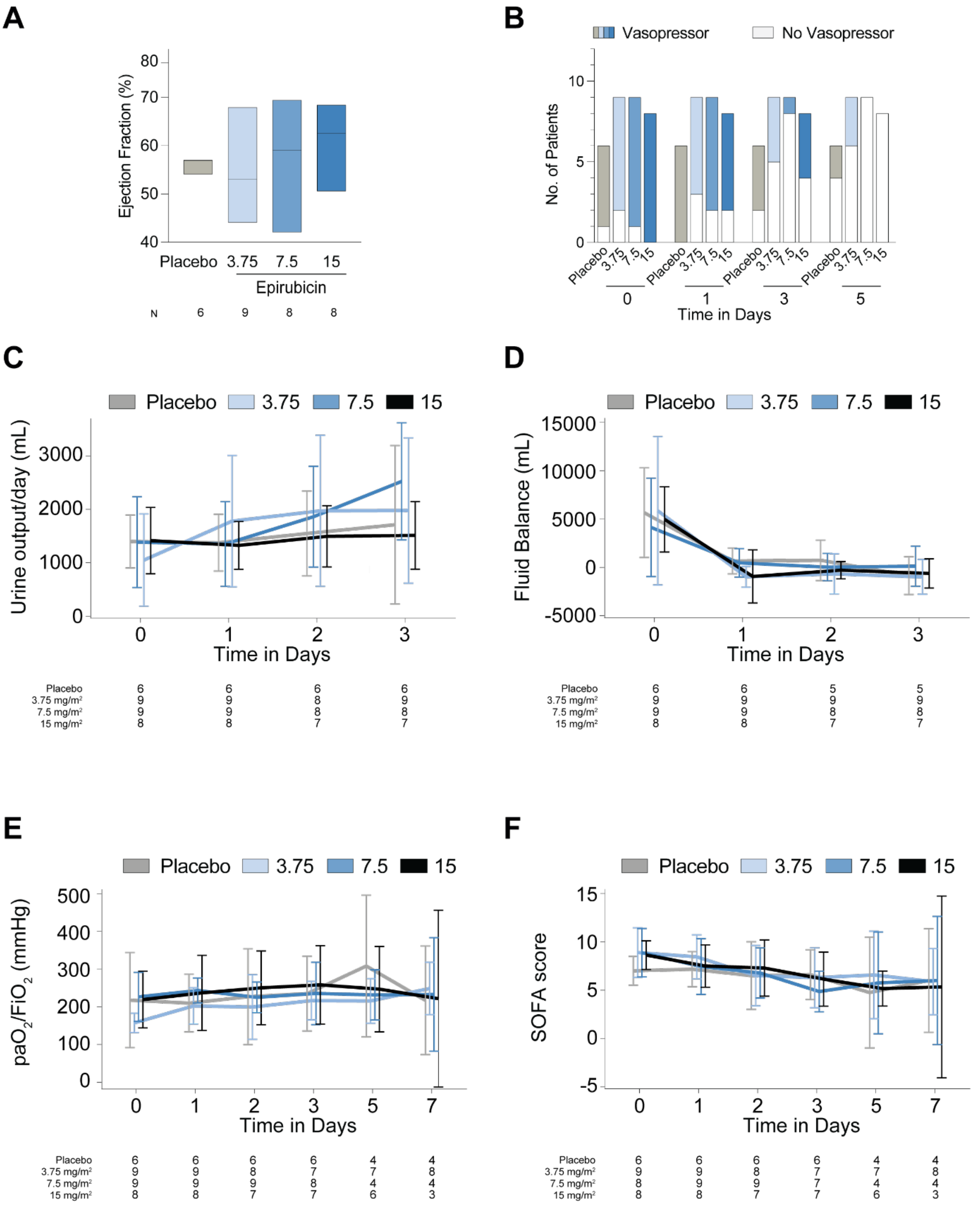


**Suppl.Figure 3: A**) Left ventricular ejection fraction assessed by TTE 7 days after intervention. Data are shown as median and IQR. **B**) Number of patients with or without vasopressors; Time course of urine-output **(C)**, fluid balance **(D)**, ratio of $paO_2/FiO_2$, **(E)** over time and SOFA score **(F)**. Data are shown as median and IQR. **F**) Number of patients with or without successful PCT reduction, **G**) plasma Interleukin-6 levels at baseline and 24 hours after placebo or epirubicin infusion.

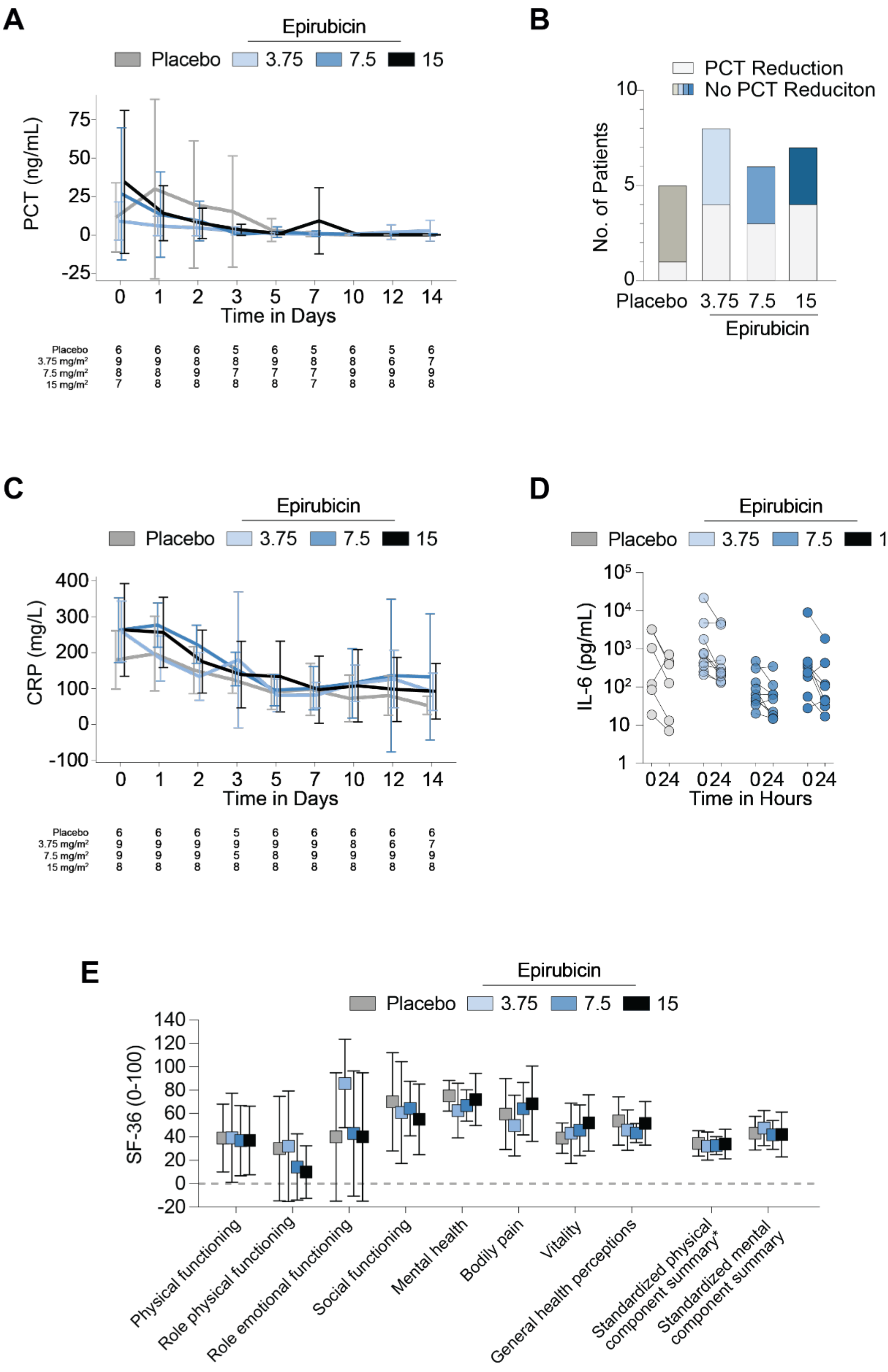


**Suppl.Figure 4: A**) Time course of PCT plasma concentrations; **B)** number of patients with and without successful PCT reduction**. C**) Time course of CRP plasma concentrations; **D**) plasma Interleukin-6 levels at baseline and 24 hours after placebo or epirubicin infusion. Data are shown as median and IQR. **E**) Assessment of quality of life using the SF-36 questionnaire at day 90. Data are shown as mean and SD.

#### 2.3 Supplementary Tables

| **Inclusion Criteria** |
|---|
| Patients $\geq$18 years admitted to the ICU/IMC with sepsis or septic shock. |
| Sepsis diagnosis within 24 hrs prior to screening regardless of site of infection. * |
| Informed consent of patient or their legal representative or if not possible a statement by an independent physician. |
| **Exclusion criteria** |
| Leukopenia/Neutropenia/Thrombocytopenia-prior or upon inclusion (Leukocyte Count <4,000/µL; Neutrophil/Thrombocyte Count below Lower Limit of Normal). |
| Weight >135 kg/BMI >45. |
| Active neoplasia.** |
| History of chemotherapy. |
| Hypersensitivity to epirubicin. |
| History of bone marrow or solid organ transplantation. |
| Immunosuppressive therapy. |
| Acute severe infection within 4 weeks prior to admission (Hospitalization for an infection or in case of hospital acquired infection transfer to a higher level of care due to the infection). |
| Chronic infection. |
| Cardiomyopathy with a documented ejection fraction <30% or ICD implantation. |
| Acute liver failure following the European Association for the Study of the Liver definition as International Normalized Ratio (INR) >1.5 and elevation of transaminases > 3 times of the upper normal limit. |
| Pregnancy during all trimester/breast-feeding. |
| Chronic mechanical ventilation dependency. |
| Cystic fibrosis. |
| Concomitant medication with Verapamil or Cimetidine. |
| Prior enrollment in this study. |
| Participation in another clinical intervention trial. |

**Suppl. Table 1**. Inclusion and exclusion criteria of the EPOS-1 trial.
* changed to 48 hours with $4^{th}$ protocol amendment. ** omitted with $4^{th}$ protocol amendment.

| | **Neutropenia** (acute neutrophil count) | **Thrombocytopenia** (platelets) |
|---|---:|---:|
| **Grade 1** | < Lower limit of normal-1,500/µL | < Lower limit of normal-75,000/µL |
| **Grade 2** | < 1,500-1,000/µL | < 75,000-50,000/µL |
| **Grade 3** | < 1,000-500/µL | < 50,000-25,000/µL |
| **Grade 4** | < 500/µL | < 25,000/µL |

**Suppl. Table 2**: Grading of neutropenia and thrombocytopenia (following [1]).

| | Exact Conditional Tests | | | |
|---|---|---|---|---|
| **Effect** | **Test** | **Statistic** | **Exact (p-Value)** | **Mid (p-Value)** |
| Group | Score | 1.9825 | 0.6210 | 0.6053 |
| | Probability | 0.0315 | 0.6522 | 0.6365 |
| Time | Score | 3.2759 | 0.2630 | 0.2440 |
| | Probability | 0.0379 | 0.3190 | 0.3000 |
| | **Exact Odds Rations** | | | |
| **Parameter** | **Estimate** | **95% Confidence Limits** | | **p-Value** |
| 3.75 mg/m$^2$Epirubicin | 0.622 | 0.007 to 52.321 | | 1.0000 |
| 7.5 mg/m$^2$ Epirubicin | 0.562 | 0.007 to 47.204 | | 1.0000 |
| 15 mg/m$^2$ Epirubicin | 2.058 | 0.144 to 119.425 | | 0.9673 |
| Time 28-Days | 1.051 | 0.013 to 84.856 | | 1.0000 |
| Time 90-Days | 4.502 | 0.419 to 231.158 | | 0.3277 |

**Suppl. Table 3**: Logistic regression analysis of mortality using exact conditional test and exact odds ratios.

| | Placebo | Phase I: 3.75 mg/m² | Phase II: 7.5 mg/m² | Phase III: 15 mg/m² | Epirubicin all patients |
|---|---|---|---|---|---|
| **Mortality, No (%)** | **N (%)** | **N (%)** | **N (%)** | **N (%)** | **N (%)** |
| Day 0 to 14 | 0/6 (0) | 1/9 (11.1) | 0/9 (0) | 0/8 (0) | 1/26 (3.8) |
| Day 15 to 28 | 1/6 (16.7) | 0/8 (0) | 0/9 (0) | 0/8 (0) | 0/25 (3.8) |
| Day 29- 90 | 0/5 (0) | 0/8 (0) | 1/9 (11.1) | 0/8 (0) | 4/25 (16.0) |
| **Total** | 1/6 (16.7) | 1/9 (11.1) | 1/9 (11.1) | 3/8 (37.5) | 5/26 (19.2) |

**Suppl. Table 4**: Overall 14-; 28- and 90-day mortality in placebo and intervention groups.

| Treatment | Day of death [days after infusion] | Cause of Death |
|---|---|---|
| Placebo | 20 | Multi organ failure |
| Phase I: 3.75 mg/m² | 10 | Respiratory failure |
| Phase II: 7.5 mg/m² | 35 | Aortic aneurysm |
| Phase III: 15 mg/m² | 47 | Chronic liver failure |
| Phase III: 15 mg/m² | 77 | Unknown |
| Phase III: 15 mg/m² | 52 | Unknown |

**Suppl. Table 5**: Reason of death in placebo and intervention groups.

| | | Placebo (all 3 phases) | Phase I 3.75 mg/m² | Phase II 7.5 mg/m² | Phase III 15 mg/m² | Epirubicin all |
|---|---|---|---|---|---|---|
| **Property** | **Answer** | **N (%)** | **N (%)** | **N (%)** | **N (%)** | **N (%)** |
| Adverse Event Severity | mild | 8 (20.0) | 9 (22.5) | 6 (24.0) | 14 (23.0) | 29 (23.0) |
| | moderate | 20 (50.0) | 15 (37.5) | 5 (20.0) | 31 (50.8) | 51 (40.5) |
| | severe | 12 (30.0) | 16 (40.0) | 14 (56.0) | 16 (26.2) | 46 (36.5) |
| Adverse Event Relationship to IMP | not related | 39 (97.5) | 33 (82.5) | 23 (92.0) | 56 (91.8) | 112 (88.9) |
| | related | 1 (2.5) | 7 (17.5) | 2 (8.0) | 5 (8.2) | 14 (11.1) |
| Adverse Event Relationship to relevant concomitant medication | not related | 40 (100.0) | 30 (75.0) | 21 (84.0) | 58 (95.1) | 109 (86.5) |
| | related | 0 | 10 (25.0) | 4 (16.0) | 3 (4.9) | 17 (13.5) |
| Adverse Event Relationship to underlying disease | not related | 18 (45.0) | 11 (27.5) | 3 (12.0) | 23 (37.7) | 37 (29.4) |
| | related | 22 (55.0) | 29 (72.5) | 22 (88.0) | 38 (62.3) | 89 (70.6) |
| Adverse Event Outcome | resolved | 28 (70.0) | 27 (67.5) | 17 (68.0) | 46 (75.4) | 90 (71.4) |
| | resolved with sequels | 3 (7.5) | 4 (10.0) | 3 (12.0) | 2 (3.3) | 9 (7.1) |
| | ongoing | 4 (10.0) | 8 (20.0) | 0 | 3 (4.9) | 11 (8.7) |
| | ongoing until death | 3 (7.5) | 0 | 4 (16.0) | 7 (11.5) | 11 (8.7) |
| | fatal | 1 (2.5) | 1 (2.5) | 1 (4.0) | 3 (4.9) | 5 (4.0) |
| | unknown | 1 (2.5) | 0 | 0 | 0 | 0 |

**Suppl. Table 6**: Severity of AEs (mild/moderate/severe), adverse event outcome (resolved/ongoing/fatal) and relation to IMP, relevant concomitant medication and underlying disease

| | | Placebo (all 3 phases) | Phase I 3.75 mg/m² | Phase II 7.5 mg/m² | Phase III 15 mg/m² | Epirubicin all |
|---|---|---|---|---|---|---|
| **Property** | **Answer** | **N (%)** | **N (%)** | **N (%)** | **N (%)** | **N (%)** |
| Adverse Event SAE | no | 30 (75.0) | 25 (62.5) | 13 (52.0) | 49 (80.3) | 87 (69.0) |
| | yes | 10 (25.0) | 1 (37.5) | 12 (48.0) | 12 (19.7) | 39 (31.0) |

**Suppl. Table 7**: Numbers and frequency of Serious Adverse Events in placebo and intervention group.
